# Reproducible profiling of the gut microbiota using surplus clinical Faecal Immunochemical Test (FIT) samples

**DOI:** 10.1101/2025.07.04.25330898

**Authors:** Merel A van den Haak, Jakub T Zbikowski, Aliu Moomin, Joan Wilson, Chris Halsey, Charlie Gourley, Farhat VN Din, Stephen T McSorley, Elaina SR Collie-Duguid, Graham Horgan, Alan W Walker, Alexandra M Johnstone, Anne E Kiltie

**Affiliations:** The Rowett Institute, University of Aberdeen, UK; Aberdeen Cancer Centre, University of Aberdeen, UK; NHS Grampian Biorepository, Aberdeen Royal Infirmary, UK; School of Cancer Sciences, University of Glasgow, UK; Edinburgh Cancer Research, University of Edinburgh, UK; Centre for Genome-Enabled Biology and Medicine, University of Aberdeen, UK; Biomathematics & Statistics Scotland, Foresterhill, Aberdeen, UK

**Keywords:** qFIT, Quantitative Faecal Immunochemical Test, gut microbiota, bowel screening, 16S rRNA gene sequencing, EXTEL HEMO-AUTO MC

## Abstract

**Background:** Numerous countries use the EXTEL HEMO-AUTO MC Quantitative Faecal Immunochemical Test (qFIT) to screen for faecal haemoglobin. We aimed to determine if bacterial 16S rRNA gene sequencing results from the leftover qFIT cassettes would be stable over time and comparable with larger volume faecal collection protocols.

**Methods:** Four qFIT probe samples were taken from each of sixteen fresh healthy volunteer stool samples and sequencing results were compared after 0, 4, 7 and 14 days to provide a baseline control and mimic postage and sample processing conditions in cancer screening programmes. qFIT results were then compared to those of standard laboratory processing of larger whole-stool samples. DNA was extracted from 100 NHS surplus qFIT samples from symptomatic patients reporting rectal bleeding and quantified to assess suitability for 16S rRNA gene sequencing.

**Results:** Bacterial composition and diversity from healthy volunteer qFITs remained stable over 14 days with only minor differences compared to baseline (day 0) and larger stool control samples; at least 75% of the symptomatic qFITs yielded sufficient DNA for 16S rRNA gene sequencing.

**Conclusion:** qFIT samples were almost identical to control samples and stable over 14 days, allowing them to be used for large-scale low-cost population-based intestinal microbiota studies.

**Clinical Trial Registration:** The study was registered on clinicaltrials.gov (NCT06100549).

**Data summary:** The datasets generated from human samples supporting the conclusions of this article are available in the Figshare repository: https://figshare.com/s/3e75fcba093f13750402. The raw sequencing datasets used for this study have been deposited in the NCBI Short Read Archive, under accession number PRJNA1268008 https://www.ncbi.nlm.nih.gov/bioproject/PRJNA1268008.

**Impact statement:** The quantitative Faecal Immunochemical Test (qFIT) is now widely used in colorectal cancer screening and symptomatic testing to detect faecal haemoglobin, with millions of samples tested annually worldwide. As only microlitres of the sample-containing buffer are required for haemoglobin testing, this provides a potentially very valuable, otherwise wasted resource of surplus samples, which could be used to analyse the gut microbiota composition by 16S rRNA gene sequencing or metagenomic methods.

We have demonstrated that the EXTEL HEMO-AUTO MC collection picker, widely used for screening and symptomatic testing, and which uses only 2 mg of faeces, yields sufficient DNA for microbiota sequencing (with stability over 14 days) and 16S rRNA gene sequencing results were generally comparable to those from larger, conventionally collected faecal samples.

An ability to reliably measure gut microbiota composition in surplus qFIT samples, with associated data linkage, could lead to large-scale, low-cost, population-based studies, including longitudinal studies from ages 50 to 74 years. Such an approach would provide unparalleled statistical power to assess links between the gut microbiota and some of the most prevalent causes of poor health, including cardiovascular disease, chronic inflammatory disease and diabetes, as well as providing vital insights into the gut microbiota during cancer development and treatment response.

**Repositories:** The raw sequencing datasets used for this study have been deposited in the NCBI Short Read Archive, under accession number PRJNA1268008.

## Introduction

Bowel screening programmes, active in many countries around the world, use qFIT (quantitative Faecal Immunochemical Testing) to screen a small faecal sample for the presence of haemoglobin, followed by a colonoscopy when the test is positive, thereby improving early detection of colorectal cancer. For example, approximately 0.5 million samples are obtained in the Scottish National Bowel screening programme (SNBSP) per year. Faecal haemoglobin sampling and measurement is therefore relevant to the majority of the 44,000 patients diagnosed with CRC each year in the UK (1,2). This test, the qFIT, is also recommended for testing patients reporting symptoms considered to be concerning for colorectal cancer (CRC) to their General Practitioner (GP) (3). In numerous sites throughout Scotland, England, Ireland, Wales, Japan and Taiwan, FIT testing uses the EXTEL HEMO-AUTO MC/ HM-JACKarc system (James Rae, Alpha Laboratories Ltd, personal communication), while some other countries use the OC-Sensor (Eiken, Japan) or FOB Gold (Sysmex, UK) FIT systems.

The gut microbiota, encompassing all microorganisms living in the gastrointestinal tract, plays important roles in human health. Perturbations in microbiota composition have been linked to cardiovascular disease (4), inflammatory bowel disease (5) and the development and progression of colorectal cancer (6). One of the most widely used methods to study gut microbiota composition and diversity is 16S rRNA gene sequencing. The 16S rRNA gene is present in all bacteria. Conserved regions in the 16S rRNA gene allow for universal amplification of bacterial DNA, and variable regions are utilised to distinguish between different bacterial taxa (7,8).

If faecal samples already collected for bowel cancer screening or symptomatic testing reliably represent the gut microbiota, then such samples could potentially be used to provide large-scale low-cost population-based faecal gut microbiota data. At a population level, this could be linked to ‘big data’ within trusted research environments, to provide secure access to local health data for research (9–12). With the possibility of longitudinal sampling and data linkage, the data obtained using routine qFIT faecal sampling could be used to explore associations between the gut microbiota, ageing and diseases including cancer (13) in large, condition-relevant cohorts. Such an approach would provide unparalleled statistical power to assess links between the gut microbiota and some of the most prevalent causes of poor health, including cardiovascular disease, chronic inflammatory disease and diabetes, as well as providing vital insights into the gut microbiota during cancer development and treatment response. Stool sampling using FIT has been reported to be widely acceptable and feasible in patients with lower GI symptoms in primary care (14) and is familiar to clinical teams. This, coupled with existing embedded pathways and protocols for requesting, stocking, collection, transport, processing and storage of FITs, provides a significant advantage over fresh stool for both sampling at scale and repeat sampling in large patient populations.

Several studies have already investigated the possibility of using varying FIT approaches for gut microbiota research, for example the OC-sensor (Eiken, Japan) FIT test, the FOB-Gold FIT (Sentinel, Italy) and the QuickRead go iFOB FIT tubes (Aidain, Espoo, Finland) with promising results, although storage temperatures and buffer composition could have an influence on sample stability over time (15,16,18–20). However, each of these FITs collect 10 mg of faeces in 1.7 to 2 ml of buffer, in contrast to the smaller 2 mg amount of faeces in 2 ml buffer for the EXTEL HEMO-AUTO MC collection picker. While microbial profiles of OC-sensor, QuickRead iFOB and FOB-Gold faecal samples have been studied, to our knowledge, no microbial research has been done on the EXTEL HEMO-AUTO MC qFIT collection device, which is an important knowledge gap given the smaller amount of faeces collected and its widespread use in various countries for routine screening programmes.

Our aim was to test the hypotheses that a) the small amount of faecal material contained within the EXTEL HEMO-AUTO MC qFIT cartridges is sufficient to perform 16S rRNA gene sequencing, b) bacterial proportional abundance and diversity, found in the qFITs, would be stable over time (0, 4, 7 and 14 days) commensurate with clinical sample handling, and c) the microbial community found in the qFIT is comparable with larger volume faecal collection protocols. We further tested whether patients’ routine use of the test kits results in sufficient DNA yield for microbiota sequencing by measuring DNA yield from 100 symptomatic qFITs and comparing this to the minimum DNA yield that resulted in successful 16S rRNA gene sequencing in this study.

## Methods

### Study participants

Two frozen stool samples from patients diagnosed with prostate cancer, stored at −70°C (18 months for sample 1 and 20 months for sample 2) and previously used for another ethically approved study (26), were obtained from the NHS Grampian Biorepository, part of the Research Scotland Biorepository, for assay optimisation tests (27) (from here on referred to as “sample set one”).

Sample set two consisted of sixteen healthy participants, six male and 10 female, mean volunteer age 28.9 years (range: 19 to 57), mean body mass index (BMI) 24.1 kg/m^2^ (range 20.4 to 29.1). Volunteers had no history of cardiovascular disease, diabetes, bowel disease, autoimmune disorders or cancer. Participants had not taken any antibiotics three months before the faecal sample collection. All participants provided written informed consent before entering the study. The study protocol was approved by the Rowett Institute Ethical Committee and the NHS Grampian Biorepository (TR000330), and the study was registered on clinicaltrials.gov (NCT06100549).

For sample set three, following ethical approval, NHS Grampian Biorepository provided 100 anonymous surplus qFIT samples from symptomatic patients who reported rectal bleeding and were therefore asked to do a qFIT test by their GP. These samples were received from the Biochemistry laboratory in Aberdeen Royal Infirmary, after 3-6 days of sample storage at 4°C following haemoglobin testing.

### Sample collection for determining optimal qFIT buffer volume for 16S rRNA gene sequencing

The qFIT probes (EXTEL HEMO-AUTO MC collection picker, Minaris Medical, now Canon Medical Diagnostics Ltd.), generously gifted to us by Alpha Laboratories Ltd, are designed to collect 2 mg of stool (excess stool is removed on probe insertion) into the collection device holding 2 ml of buffer. Haemoglobin testing of the qFIT sample using the HM-JACKarc analyser (Hitachi Chemical Diagnostics Systems, Japan) only requires 6 µl of the 2 ml buffer/faeces suspension, after which the qFIT cartridge is stored at the NHS laboratory at 4°C to allow for a repeat screening in case of failure, before being discarded. Symptomatic samples are stored for a week post analysis at 4°C (Catriona Cameron, NHS Grampian, personal communication).

Initial testing was performed on sample set one, containing frozen faecal samples obtained for a previous study (26) from two patients diagnosed with prostate cancer (Figure 1A).

**Figure 1.**
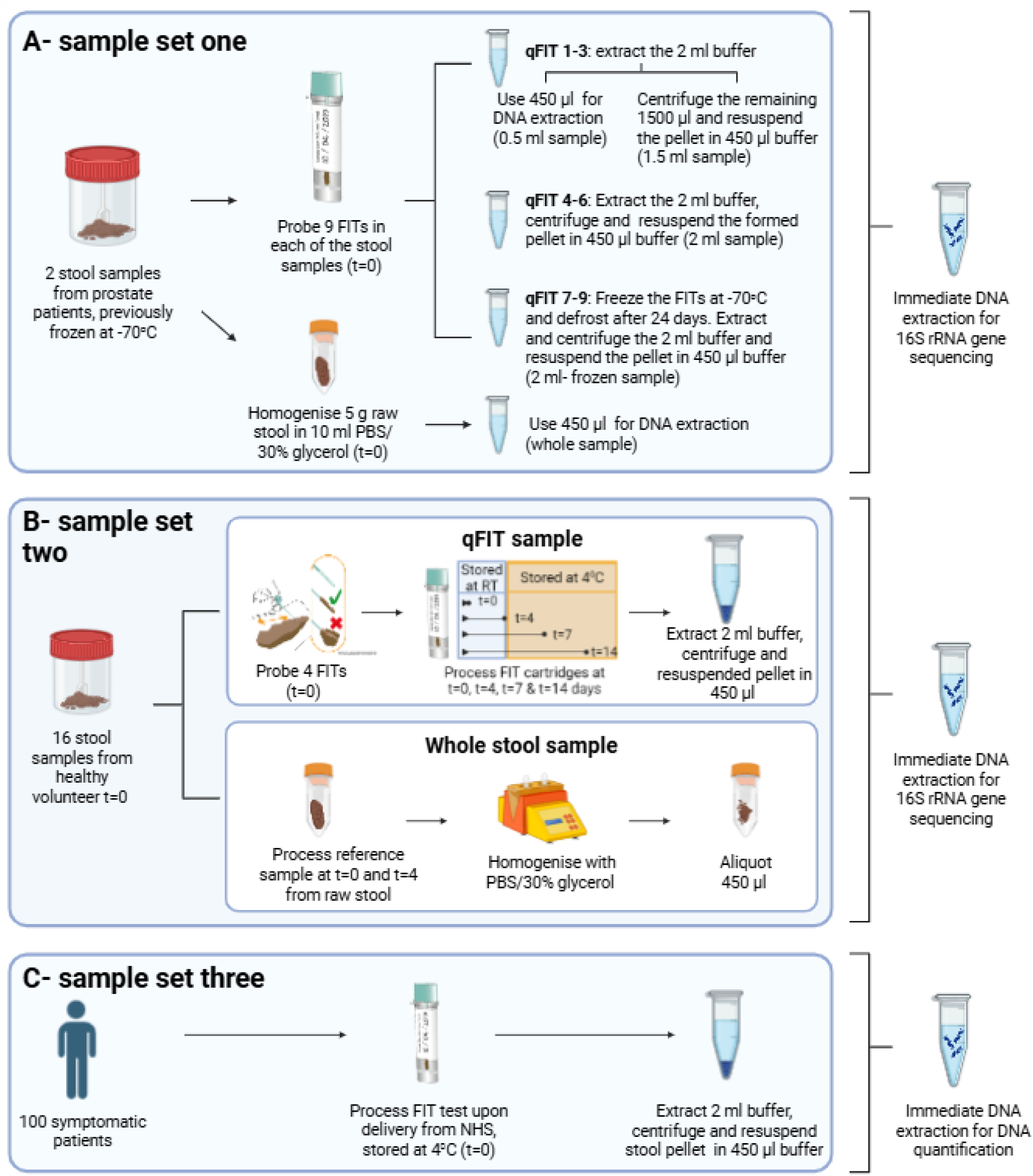
Schematic showing sample processing steps for each participant group. A) For sample set one, two stool samples were obtained from a previous study for assay optimisation. B) For sample set two, DNA was extracted from four qFIT cartridges and two whole stool-derived samples per healthy volunteer (n=16). The 16S rRNA gene sequencing results from the qFIT samples were compared to the whole stool samples. C) For sample set three, DNA was extracted from 100 anonymised qFIT samples from symptomatic patients, solely to assess DNA yield. The figure was created with BioRender.com.

Due to the small faecal sample size in the qFIT cassette, this pilot sample set was used to determine whether DNA yield could be optimised to aid 16S rRNA gene sequencing, and to establish if the entire 2 ml buffer volume was required to obtain satisfactory results. To do this, nine separate qFIT probes were inserted in each sample and placed in their respective cassettes. From qFIT 1-3, 450 µl of the 2 ml qFIT sample containing faeces was directly used for DNA extraction. The residual 1500 µl of each of these samples was then centrifuged at 3000 rpm for 15 minutes until pellet formation and the resulting faecal pellet was resuspended in 450 µl of the removed supernatant before DNA extraction. This resulted in the 450 µl final volume containing the faecal pellet originally suspended in the 1500 µl, potentially increasing DNA yield three-fold. Using the same principle, the entire 2 ml qFIT sample buffer volume from qFITs 4-6 was extracted from the cartridge on the same day and centrifuged, after which the formed pellet was resuspended in 450 µl supernatant and used for immediate DNA extraction. qFITs 7-9 were frozen at −70°C for 24 days and processed using the same process as qFITs 4-6. These samples were then compared to the whole stool sample, comprising of 5 g of raw faecal sample and 10 ml of PBS/30% glycerol buffer; 450 µl of this homogenised sample was used for DNA extraction, and the results compared to the qFIT samples.

### Healthy volunteer faecal sample collection and processing

For sample set two, sixteen healthy participants were instructed to collect a 17-18 ml stool sample into a Sterilin 30 ml stool sample collection tube (Alpha Laboratories, UK) using a Fe-Col® faecal collection paper (Alpha Laboratories, UK), and to deliver the sample to the lab within 18 hours. The samples were kept cool in cool bags containing frozen freezer blocks, while ensuring they did not freeze, and sample processing commenced immediately upon delivery to the Rowett Institute.

Four different qFIT probes were inserted multiple times into each sample until both serrations of the sample collection stick were appropriately filled (Figure 1B). Probe one was immediately processed for DNA extraction upon delivery of the sample (t=0). The remaining qFIT cartridges were stored for times and temperatures commensurate with postage and sample processing in the Scottish National Bowel Screening programme, corresponding to a time course of 4 days to 2 weeks. Probe 2 was stored at room temperature and DNA extracted on t=4 days to mimic postage/shipping conditions of the qFIT to the lab. As testing laboratories store the qFIT cartridges at 4°C, after storage at RT for 4 days, probes 3 and 4 were then stored at 4°C. DNA was extracted at t=7 days and t=14 days post-sample collection respectively. The cartridges were vortexed for 2 minutes to ensure proper homogenisation before processing. Then the entire sample was removed from the qFIT cartridges using sterile Pasteur pipettes, centrifuged at 13,000 rpm for 15 minutes, the supernatant removed, and the pellet resuspended in 450 µl of the removed supernatant.

The qFIT samples from each volunteer were compared to their whole faecal sample, collected and processed using a method which we have previously demonstrated to be acceptable in a clinical setting (28), to determine comparability of the sequencing data. A homogenised, 5 g aliquot of raw stool was taken from the main faecal sample, suspended in 10 ml PBS solution with 30% glycerol and homogenised using a gentleMACS™ Dissociator (Miltenyi Biotec Ltd., UK). DNA was immediately extracted from a 450 µl aliquot. This was repeated at t=4 using another 5 g aliquot of the raw stool sample that had been stored at 4°C (Figure 1C).

### Symptomatic qFIT sample processing

For sample set three, containing the 100 anonymous samples obtained from symptomatic patients (Figure 1C), 2 ml qFIT buffer containing faecal sample was extracted from the cartridges, centrifuged and the supernatant removed at t=0, when the samples were received in the lab. The pellet was resuspended in 450 µl of the removed qFIT sample supernatant prior to DNA extraction. Before arrival to our lab, the samples had been stored at 4°C at the UK National Health Service clinical lab for haemoglobin testing.

### DNA extraction

The FastDNA SPIN Kit for Soil (MP Biomedicals, UK) was used for all DNA extractions (29). This included 26 samples for sample set one, 96 samples for sample set two and 100 samples for sample set three. Processing followed the manufacturer’s instructions with following minor adaptations: resuspended qFIT samples (450 µl) and homogenised faecal matter from the whole stool-derived samples (450 µl) were placed in individual, separate lysing matrix E tubes, and 800 µl sodium phosphate buffer and 122 µl MT buffer were added to each tube. 450 µl of liquid sample was used, and 1 ml of Binding Matrix Solution supernatant was discarded. DNA was quantified using a Qubit 4 Fluorometer (Invitrogen, UK) using the standard protocol. Extracted DNA was stored at −20°C.

### 16S rRNA gene sequencing

16S rRNA gene sequencing on sample set one and two was carried out at the Centre for Genome-Enabled Biology (CGEBM), University of Aberdeen. Three qFIT blanks were included, as were two PCR negative (water in place of gDNA) (n=3) and three positive (ATCC MSA-1003 mock community) controls. No sequencing was carried on sample set three, as its sole purpose was to show that samples taken by the general public (rather than in a controlled laboratory environment) contained a suitable amount of DNA for 16S rRNA gene sequencing, at a threshold that was already shown to be effective with sample sets one and two.

The 16S rRNA gene variable region V1-V2 was used for bacterial community profiling of the faecal samples as described previously (28). Briefly, Illumina compatible libraries were prepared using 2-step PCR with 2.5 µl of input gDNA or study negative control (PBS/ 30% glycerol buffer in place of sample) per sample, 200 nM each of forward and reverse PCR primers and 1x Kapa HotStart ReadyMix (Roche) in a reaction volume of 25 µl. PCR primer sequences were 16S V1 (Forward: 5’ **TCG TCG GCA GCG TCA GAT GTG TAT AAG AGA CAG** AGM GTT YGA TYM TGG CTC AG 3’) and 16S V2 (Reverse: 5’ **GTC TCG TGG GCT CGG AGA TGT GTA TAA GAG ACA G**GC TGC CTC CCG TAG GAG T 3’), and included a specific Illumina compatible sequence (bold) that was the primer binding site in the second PCR. Plates were sealed and PCR was performed in a thermal cycler with initial denaturation at 95°C for 3 minutes, followed by 15 cycles of 95°C for 30 sec, 55°C for 30 sec and 72°C for 30 sec, and a final extension at 72°C for 5 min. Plates were held at 4°C until further processing. For each experimental sample and control, 3 PCR reactions were prepared to improve detection of rarer taxa. Following PCR, PCR reactions were combined per sample and purified using SPRI magnetic beads (Beckman-Coulter, CA), according to the manufacturer’s instructions using a 0.8x ratio of beads:sample. Then 5 µl of purified amplicons were used as template in the second PCR with 1x Kapa HiFi Hotstart Readymix (Roche, CH) and 5 µl each of Nextera XT Index 1 and 2 primers (Nextera XT index kit v2, Illumina, CA), in a reaction volume of 50 µl. Plates were sealed and PCR was performed in a thermal cycler with initial denaturation at 95°C for 3 minutes, followed by 8 cycles of 95°C for 30 sec, 55°C for 30 sec and 72°C for 30 sec, and a final extension at 72°C for 5 min, then held at 4°C. Final libraries were quantified by fluorimetry (Qubit, Thermo Fisher, MA) and average library size determined on a Tapestation 4200 (Agilent Technologies, CA), according to the manufacturer’s instructions, to calculate molarity. All samples were equimolar pooled and sequenced on an Illumina MiSeq with 300 base pair (bp) paired end reads. Base calling and demultiplexing of raw data were performed in Illumina BaseSpace and these raw data fastq were used for downstream analysis.

### Bioinformatics

Illumina MiSeq sequences were analysed in mothur 1.48.0 (30) based on the mothur Miseq standard operating procedures (31). Forward and reverse reads were paired into contigs, and poor-quality sequences (<260 bp or >470 bp) were removed from the dataset, as well as all reads with any ambiguous bases and homopolymeric stretches longer than 7 bp. Unique sequences were aligned with the SILVA reference database (Release 119) and incorrectly aligned sequences were removed. The pre.cluster command was run to merge sequences with <3 base differences in order to mitigate the impact of sequencing errors, and split.abund was used to remove singletons (32). The Ribosomal Database Project (RDP) version 19 reference database was used for taxonomic classification (33). Reads were clustered into operational taxonomic units (OTUs) with 97% similarity. Subsampling was performed at 31,284 bp for sample set one, excluding one 2 ml sample from donor 1 due to low yield, resulting in an average Good’s coverage estimate of 99.6% (Supplementary Table 1).

Subsampling at 34,571 bp for sample set two resulted in the exclusion of seven low-yield samples, with a final average Good’s coverage of 99.8% (Supplementary Table 1). One of these excluded samples was a whole stool sample at t=0, the other samples were qFIT samples at t=0 (n=1), t=7 (n=2), and t=14 (n=3). None of the excluded qFIT samples were from the same volunteer.

### Statistical analysis

For sample set one, Metastats software (as implemented in mothur) was used to investigate significant differences in bacterial abundance among the four buffer volumes, and between the four buffer volumes and the whole stool-derived sample at the family, genus and OTU level, focussing on OTUs or phylotypes in the top 100 for each of the individual samples when more than 100 OTUs or phylotypes were present (34). The decision to focus on the top 100 OTUs from the entire dataset was made prior to data analysis to achieve a balance between examining a broad selection of dominant OTUs, without potentially increasing the risk of false negatives by incorporating very rare OTUs into false-discovery rate correction calculations. This balanced approach therefore allowed for assessing the consistency of qFIT based results vs processed whole stool. For sample set two, Metastats was used to investigate significant differences in bacterial proportional abundance between whole stool-derived samples t=0 and t=4, qFIT t=0 and whole stool-derived samples t=0, qFIT t=4 and whole stool-derived samples t=4, qFIT t=0 and qFIT t=4, qFIT t=0 and qFIT t=7 and qFIT t=0 and qFIT t=14, focussing on the top 100 OTUs or phylotypes present in each individual sample where more than 100 were present. All p-values were corrected with the Benjamini Hochberg method with cut-off set to p<0.05 to control for the false discovery rate (35).

Since it is best practice not to rely on just one statistical analysis tool (36), OTU’s and phylotypes were also tested within sample set one and sample set two using LEfSe (37) and DESeq2 (36) with FDR-adjustment in the software package MicrobiomeAnalyst to reduce the number of false positives, adding complementary statistical analysis to the Metastats findings (38). Using indices of alpha and beta diversity, it was established whether the bacterial composition of the gut microbiota differed between groups and over time. Where alpha diversity measures the microbial community features in individual samples, beta diversity measures the variation of species composition between samples (13). The alpha diversity for sample set one and two was assessed by calculating the observed richness, inverted Simpson diversity index and Shannon diversity index in mothur and further analysed using ANOVA in R studio (version 2025.05.1) (39) using the same grouping factors as described above. A principal Coordinate analysis (PCoA) plot was created in MicrobiomeAnalyst for sample set one using the Bray Curtis Index calculator by generating a distance matrix on the “shared” file from mothur, visualising the beta diversity, and significance was established using nonparametric analysis of molecular variance (AMOVA) in mothur. Dendograms for sample set one were generated in mothur using the tree.shared command with the Bray Curtis calculator with the Unweighted Pair Group Method with Arithmetic Mean (UPGMA) and visualised using iTOL (40). Parsimony software in mothur was used for sample set one and two to calculate whether clustering patterns of dendrograms were significantly different by monitoring shared branches on the tree (41).

Graphs were made in GraphPad (Version 5.04). Longitudinal associations between bacterial taxa (OTU, genus and family level) and sampling timepoint for the qFITs from sample set two were also analysed using MaAsLin2 in Rstudio with timepoint as a fixed effect (t=0 to t=14) and participant ID as a random effect, focussing on OTUs or phylotypes in the top 100 for each of the individual samples when more than 100 OTUs or phylotypes were present (42). A Benjamini–Hochberg false discovery rate correction was applied to the MaAsLin2 results. PERMANOVA was tested on both sample set one and two using the software package MicrobiomeAnalyst.

## Results

### Optimising the amount of qFIT faecal homogenate for 16S rRNA gene sequencing

Sample set one was used to investigate the optimal qFIT buffer/faeces suspension volume for gut microbiota analysis. No statistically significant differences were found between the gut microbiota profiles of the qFIT samples at the family, genus and OTU levels within the patients using Metastats and LEfSe analysis, with only *Porphyromonas* and *Peptococcaceae* found to differ between various sample sizes solely when using the DESeq2 statistical analysis approach (Supplementary Table 1), indicating a similar bacterial profile for all sample volumes and no effect of freezing for 24 days. Similarly, no statistically significant differences in bacterial proportional abundance were found between the qFIT samples and the whole stool sample (Figure 2A, Supplementary Figure 1A, Supplementary Table 1).

**Figure 2.**
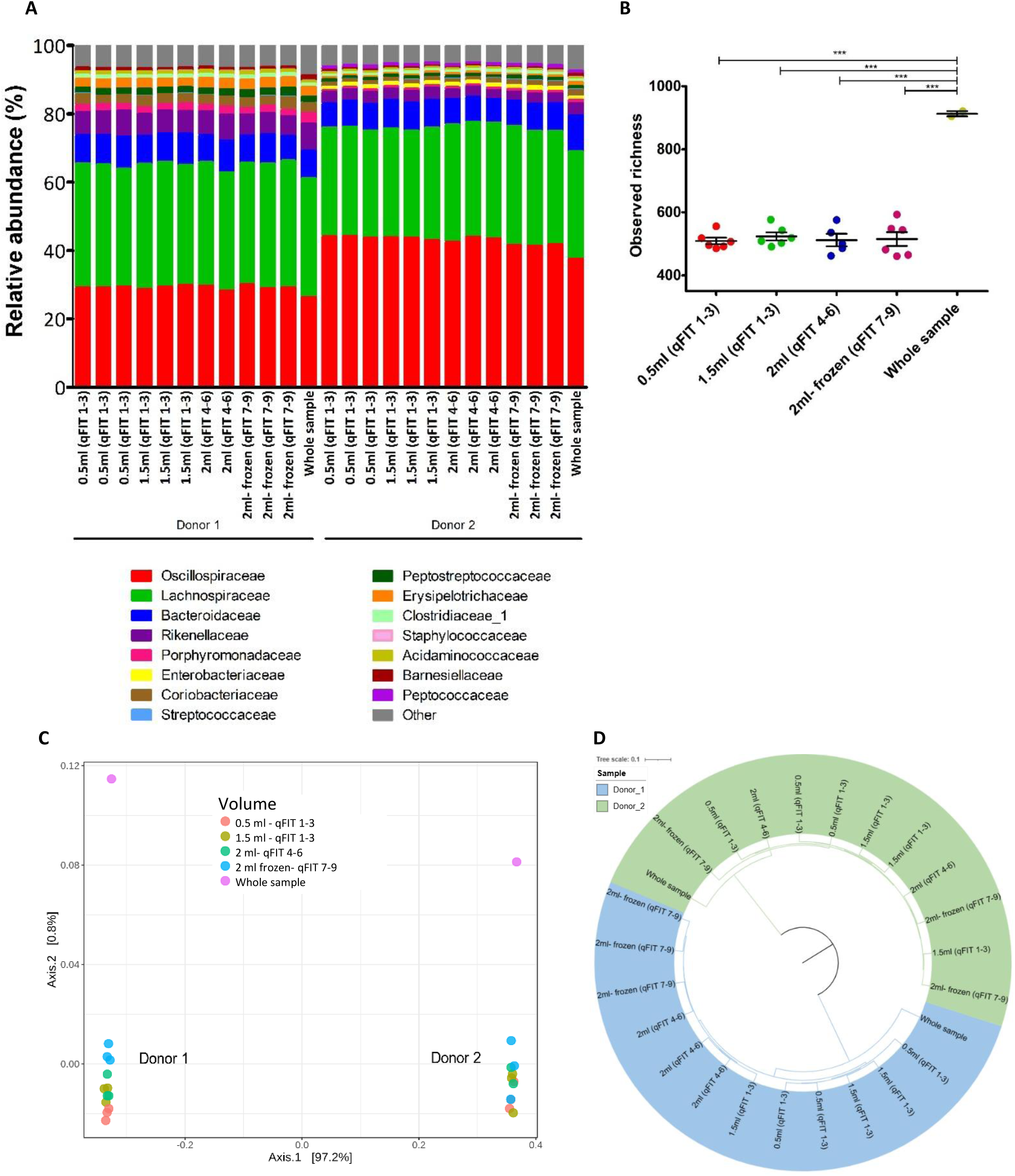
Neither the amount of qFIT buffer used for DNA extraction nor freezing influenced the 16S rRNA gene sequencing results. A) Microbiota composition, showing the 15 most proportionally abundant gut microbiota families in sample set one. One 2 ml sample from Donor 1 was excluded at the subsampling set due to low yield. Metastats with Benjamini-Hochberg correction and Benjamini-Hochberg adjusted LEfSe and DESeq2 were used to confirm the absence of statistically significant differences between the groups for the 100 most abundant OTU’s and family and genus level phylotypes; B) Observed OTU richness for each of the samples. ANOVA was conducted to compare groups for statistically significant differences; C) Principal coordinate analysis using Bray-Curtis dissimilarity. AMOVA was performed to identify significant differences across groups; D) Dendrogram made with the Unweighted Pair Group Method with Arithmetic Mean (UPGMA) method based on Bray Curtis assessment of community structure. Parsimony testing was employed to determine whether group differences were statistically significant.

There was a statistically significant increase in the number of observed OTUs present in the aliquots taken from the whole stool-derived samples versus the four qFIT buffer/faeces suspension volumes (ANOVA, p=<0.001)(Figure 2B, Supplementary Table 1), but no statistically significant differences in diversity were found using both the Inverse Simpson and Shannon diversity indices, which take into account both species richness and the evenness of species distribution (Supplementary Figure 1B and 1C, Supplementary Table 1). This indicates that although samples taken from whole stool-derived samples contained more taxa when compared to the qFIT samples, the evenness of species distribution was comparable between both sample types.

Next, beta diversity was visualised using a Bray Curtis-based Principal coordinate analysis (PCOA) plot (Figure 2C). AMOVA testing confirmed that samples clustered by donor (p=<0.001), but not by sample volume (p=0.55), indicating similar results independent of sampling method (Figure 2C). This was in agreement with PERMANOVA results, showing a significant difference between the two donors at the OTU, genus and family levels (p=0.001), but showing non-significance when comparing the different sample volumes (p=0.923 at the family level, p=0.962 at the genus and OTU levels). These results were further supported by dendrogram analysis (Figure 2D), which confirmed that samples clustered by donor (Parsimony test, p<0.001, Supplementary Table1) but not by sample volume (p=NS).

As these results showed that the volume of qFIT sample used for DNA extraction did not influence the results, the entire 2 ml qFIT buffer/faeces suspension was used for all further testing.

### Comparison of samples taken from whole stool versus qFIT faecal samples

For sample set two, DNA was extracted at t=0 and t=4 for the whole stool samples, and at t=0, t=4, t=7 and t=14 days for the four qFIT samples for each of the sixteen healthy volunteers. When comparing alpha diversity measures, it was found that the whole stool-derived samples contained a significantly higher OTU richness compared to the qFIT buffer/faeces suspension samples (ANOVA, p=<0.001) (Figure 3A, Supplementary Table 1). However, there were no statistically significantly differences between the groups when comparing the Shannon and inverse Simpson diversity indices (Figure 3B, 3C, Supplementary Table 1), indicating a similar evenness of species distribution between the whole stool-derived samples and the qFIT buffer/faecal suspensions. No statistically significant changes were found for the OTU richness, Shannon and inverse Simpson alpha diversity measures when comparing just the qFIT samples, indicating similar species distribution over time. When comparing overall microbiota structure, whole stool-derived samples and qFIT samples clustered primarily by volunteer (Figure 4), with Parsimony testing indicating no significant differences between the samples derived from whole stool and the qFIT samples at t=0 and t=4 respectively, or between the qFIT timepoints (Supplementary Table 1). PERMANOVA showed a non-significant difference when comparing the whole stool samples to the qFIT samples (p=0.074 at the family level, p=0.161 at the genus level and p=0.764 at the OTU level), and no significant differences were found between different timepoints for the whole stool samples and qFIT samples (p=0.963 at the family level and p=1 at the genus and OTU level). This indicates a comparable species composition and further strengthened the evidence for similarity between the whole stool-derived samples and the qFIT samples containing faeces suspended in buffer over time.

**Figure 3.**
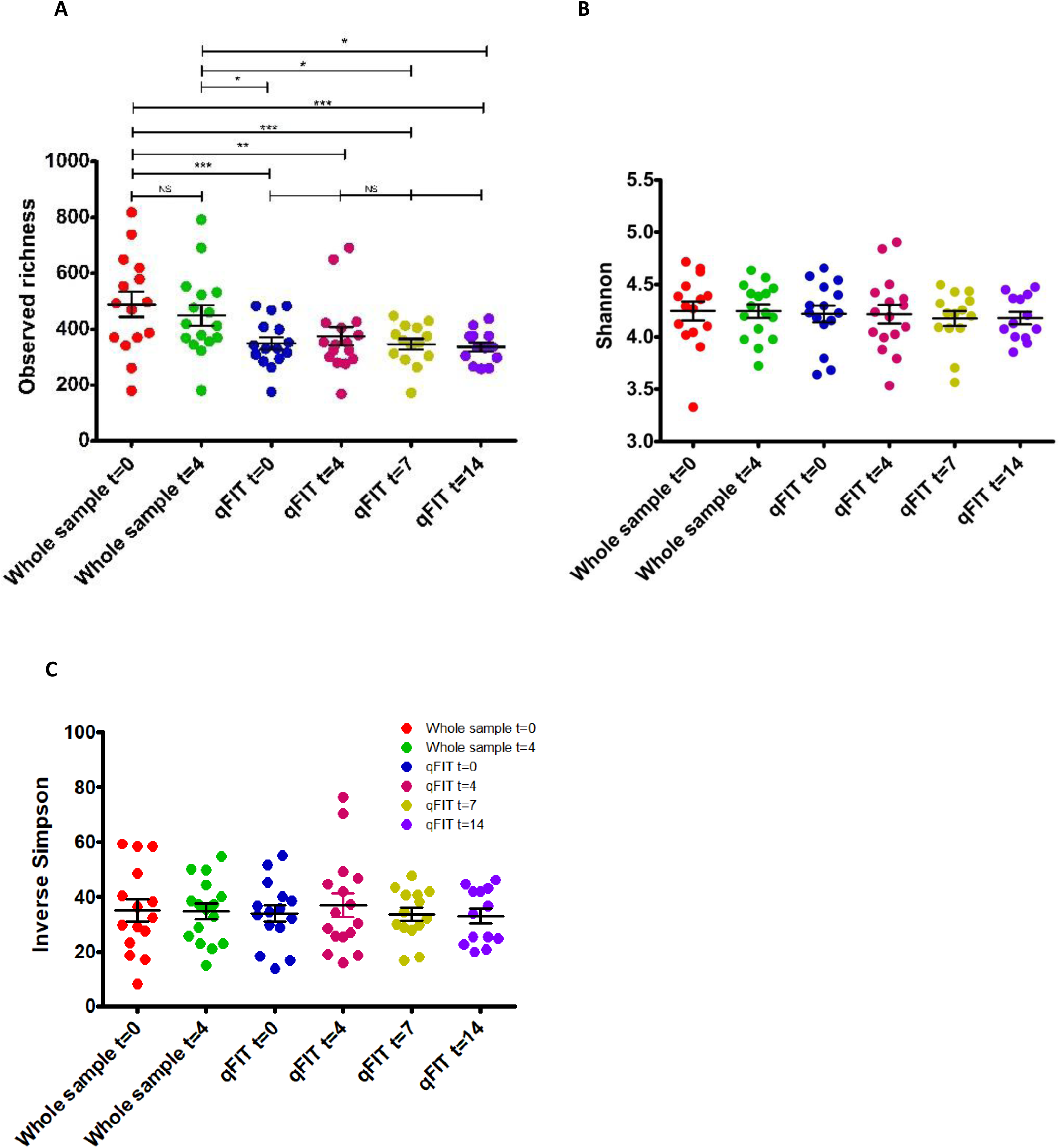
Whole stool-derived samples contained more taxa when compared to the qFIT samples, but with a comparable evenness of species distribution between both sample types. Alpha diversity measures are shown for sample set two, investigating whole stool-derived samples and qFIT samples over time. ANOVA was carried out to evaluate potential statistical differences between the groups. A) the observed OTU richness. B) Shannon diversity index. C) Inverse Simpson diversity index.

**Figure 4.**
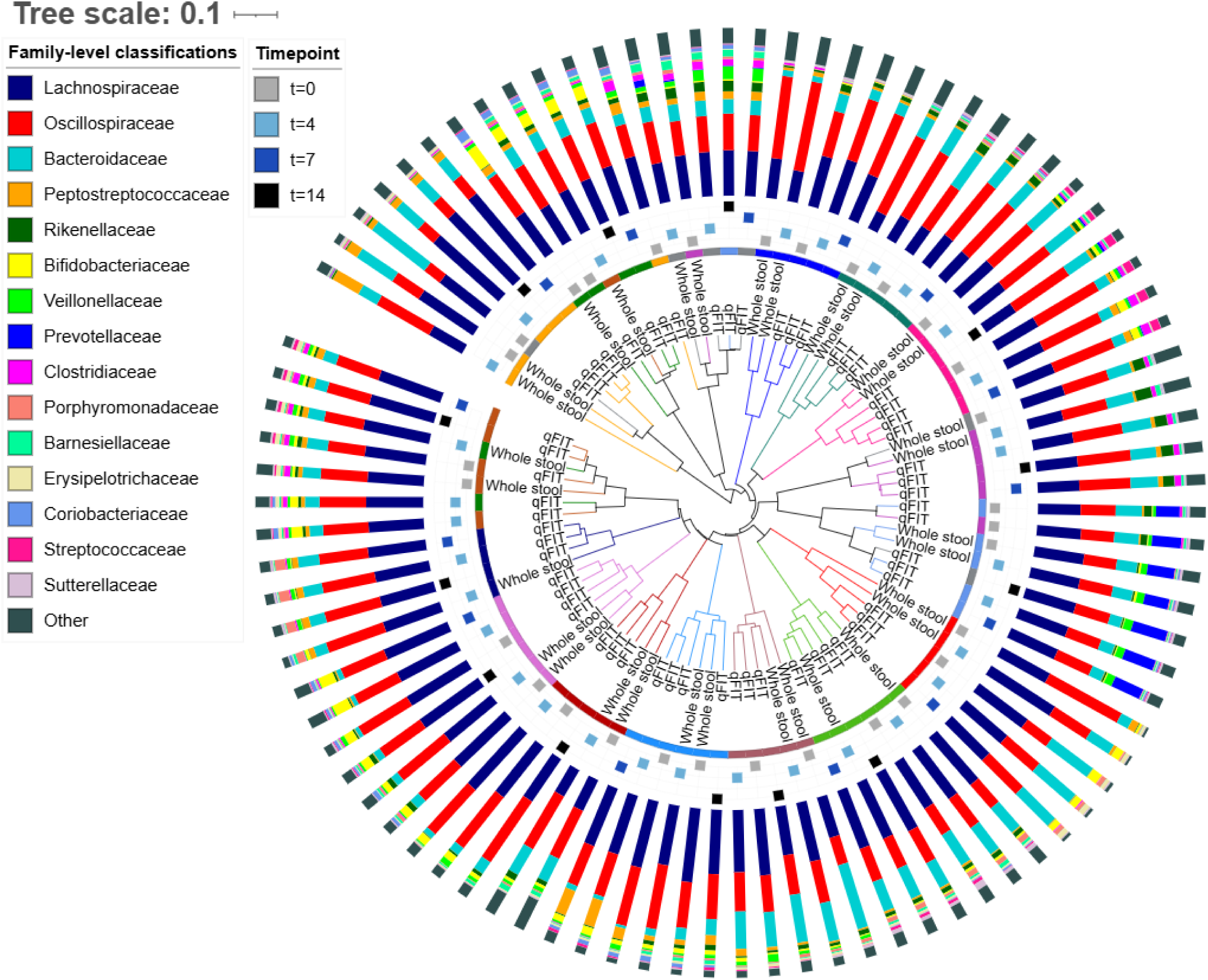
Stool-derived samples and qFIT samples clustered primarily by volunteer rather than sampling type. Parsimony testing revealed no significant differences between the whole stool samples and qFIT samples at t=0 and t=4, and qFIT samples showed no significant differences between timepoints. The dendrogram was created using the Unweighted Pair Group Method with Arithmetic Mean (UPGMA) based on the Bray Curtis-calculated dissimilarities in community structure. Individual volunteers are distinguished by colour (inner ring), and the outer ring shows the compositional profiles of the 15 most proportionally abundant gut microbiota families in sample set two.

When comparing individual taxa within the microbiota datasets, Metastats and LEfSe analysis showed no significant difference after Benjamini-Hochberg correction for multiple comparisons in either bacterial families, genera or OTUs when comparing the whole stool-derived samples at t=0 and t=4 with the qFIT samples containing faeces suspended in buffer at t=0 and t=4 respectively (Figure 5, Supplementary Figure 2, Supplementary Table 1).

**Figure 5.**
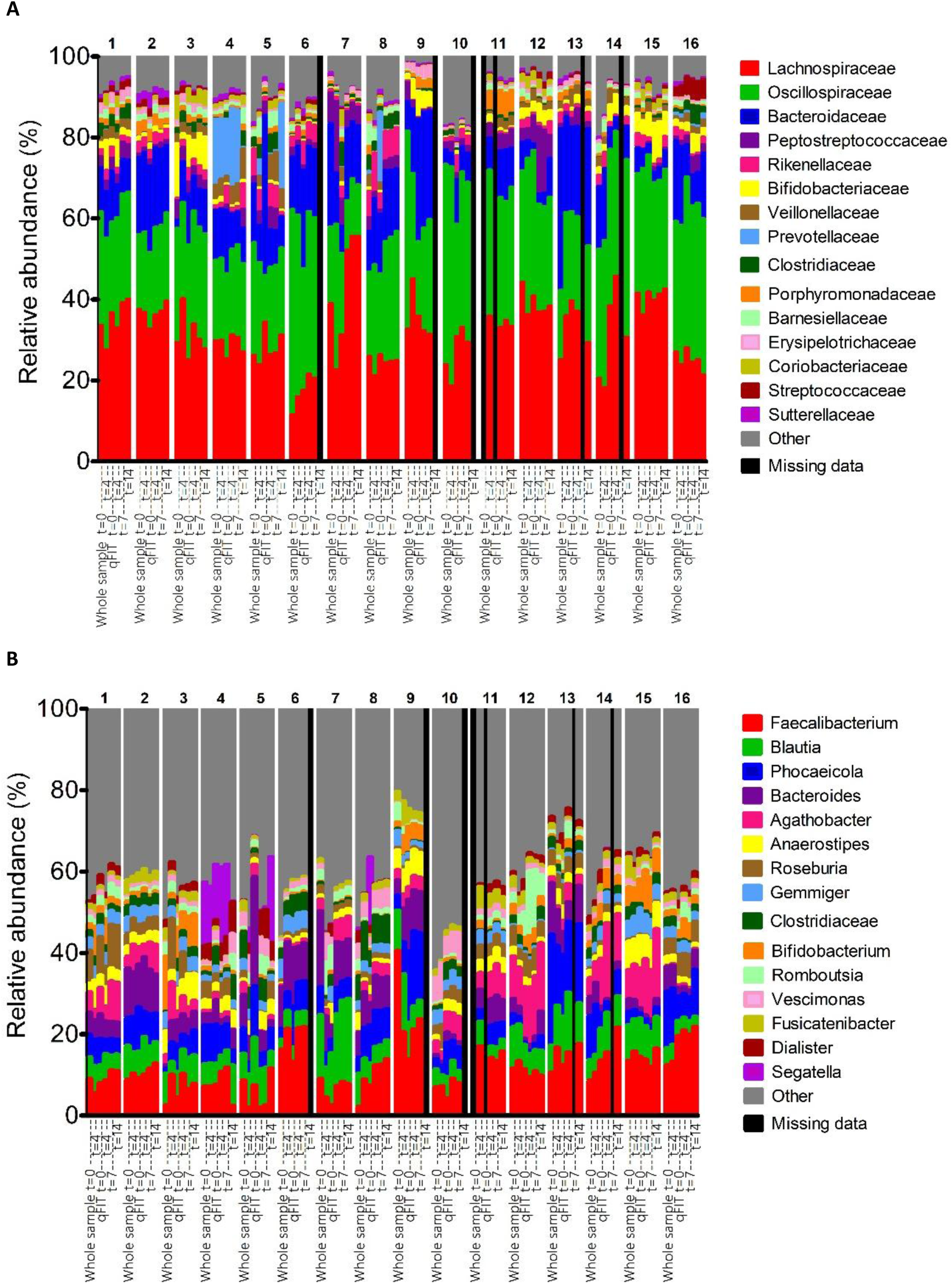
**qFIT and whole stool-derived samples showed a similar microbiota profile at the family and genus levels, with qFIT profiles remaining stable over time**. A) composition of the 15 most proportionally abundant gut microbiota families in sample set two. B) composition of the 15 most proportionally abundant gut microbiota genera. Samples are grouped by individual volunteer, shown by volunteer number, to display individual variation. Metastats on the 100 most abundant OTU’s and phylotypes with Benjamini-Hochberg correction and Benjamini-Hochberg adjusted LEfSe and DESeq2 were conducted to evaluate statistically significant differences. Longitudinal differences between the qFITs were investigated using MaAsLin2. The black vertical columns represent missing data.

DESeq2 identified *Paraprevotella* as differentially proportionally abundant between the raw stool sample and the qFIT sample at t=0 only (log_2_FC = 19.96). However, the observed differences in mean relative abundance were small (0.086% for the whole stool sample and 0.104% for the qFIT sample - Supplementary Table 1). These results therefore further indicate a similar gut microbiota profile between whole stool samples and qFIT samples.

Longitudinal MaAsLin2 analysis on the qFITs identified no significant associations between bacterial taxa at the OTU, genus and family levels and sampling timepoint (t=0 to t=14) after Benjamini-Hochberg correction for multiple comparisons (Supplementary Table 1). Similarly, no statistically significant differences in proportional bacterial abundances were consistently found at the family, genus and OTU levels with Metastats, LEfSe and DESeq2 between qFIT t=0 and t=4, qFIT t=0 and t=7 and FIT t=0 and t=14, further suggesting that the qFIT samples remained stable over time.

### Assessment of feasibility of symptomatic qFIT samples collected by members of the public

For sample set three, DNA was extracted from qFIT buffer/faecal suspension from 100 symptomatic patients, to establish whether samples taken in real-world settings by patients would result in similar DNA yields as the qFITs tested in the research laboratory. Overall, the 100 cartridges had a mean DNA yield of 1.19 ng/µl (SD 1.04 ng/µl) (Supplementary Table 1). Twenty-five samples had a DNA concentration below 0.432 ng/µl, which was the lowest DNA concentration in samples that were successfully 16S rRNA gene sequenced in this study, indicating that the DNA yield of at least 75 samples was sufficient for 16S rRNA gene sequencing.

## Discussion

Our data supports the use of the EXTEL HEMO-AUTO MC qFIT cartridge to reproducibly measure gut microbiota. This unlocks the potential of repurposing leftover qFIT bowel screening samples for microbiome research.

Initial testing on sample set one, containing two frozen stool samples from patients with prostate cancer demonstrated that the volume of qFIT buffer/faecal suspension used did not alter 16S rRNA gene sequencing results, apart from a higher observed richness being observed in the whole stool samples when compared to the qFIT samples. It is noteworthy that the frozen qFIT samples from sample set one closely resembled the other sample set one qFIT samples, indicating that samples could generate representative microbiota profiles for up to 24 days after re-freezing.

The primary aim of this study was to compare the bacterial microbiota from sample set two, containing 16 healthy participants using EXTEL HEMO-AUTO MC qFIT collection cassettes with samples collected from whole stool. The qFIT samples from this sample set were comparable to the whole stool-derived samples, except for the observed richness, likely due to improved detection of rare taxa from the larger faecal input amount (5 g versus 2 mg), demonstrating the suitability of this qFIT model for gut microbiota research. Moreover, there was no significant difference in bacterial proportional abundance, alpha diversity and beta diversity with sample storage over time, demonstrating that shipping and clinical laboratory sample handling in bowel screening programmes should not interfere with subsequent 16S rRNA gene sequence data quality, provided it is carried out within 14 days of stool collection. DESeq2 analysis identified one genus that significantly differed between the whole stool samples and qFIT samples for sample set two. However, as this result was not replicated by the other two statistical approaches that were used (Metatstats and LEfSe), and the relative abundance only modestly increased, it is clear that the qFIT samples can be considered very similar to the whole stool samples. Using sample set three, it was also found that at least 75% of samples collected by patients in the real-world setting and processed through the NHS symptomatic testing service for qFIT haemoglobin testing showed a suitable DNA yield for 16S rRNA gene sequencing.

Several studies have already investigated the possibility of using varying FIT approaches for gut microbiota research (21). Microbiota testing of DNA from faecal samples collected in the OC-sensor (Eiken, Japan) FIT test cartridges has demonstrated conserved community membership, alpha diversity and proportional abundance of genera when raw stool and FIT samples were compared (15,16), and the ten most dominant genera from the OC-sensor samples were consistent with other studies using stool samples (17). Another study examined three bacterial species and confirmed their presence in both the OC-sensor and the FOB-Gold FIT (Sentinel, Italy) and reported no loss in detection levels for up to 6 days (18). The OC-sensor cartridges also conserved bacterial diversities when compared to immediately frozen raw stool from healthy participants, providing further evidence that microbiota profiles in OC-sensor collected faecal samples and stool samples are similar (19).

We showed that the EXTEL HEMO-AUTO MC qFIT gives a similar gut microbiota composition when compared to raw stool samples. This is in agreement with previous studies (17,21,43), despite a reduction in the amount of faeces collected in the EXTEL HEMO-AUTO MC cartridge and likely differences in composition of the proprietary buffers compared to several other FIT types. The OC-sensor and QuickRead iFOB collect 10 mg faeces in 2 ml buffer (22), and the FOB-Gold uses 10 mg of stool in 1.7 ml of buffer (23), which is in contrast to the 2 mg faeces in 2 ml buffer for the EXTEL HEMO-AUTO MC collection picker. Although the buffer compositions of the FITs are proprietary, their composition likely differs, as the companies report for example different sample stabilities for the qFIT and OC-sensor (24,25). The OC-sensor guarantees haemoglobin sample stability for 7 days at RT or 28 days at 2-8°C (25). In contrast, the EXTEL HEMO-AUTO MC cartridge guarantees stable haemoglobin concentrations for 14 days at RT or 120 days at 2-8°C (24)).

Similar to our results, the OC-sensor FITs also demonstrated microbiota clustering by volunteer rather than sampling method (44,45) and displayed comparable alpha diversity and composition between the FIT and fresh whole stool-derived sample (16). However, in contrast, several other papers have found small differences in bacterial proportional abundance between stool and OS-sensor FIT samples (43,45–47), which we did not find when comparing the EXTEL HEMO-AUTO MC with the whole stool samples, potentially explained by technical or biological variation.

We showed sample stability of the EXTEL HEMO-AUTO MC over time. Similar results were found for the OC-sensor FIT and QuickRead go iFOB FIT tubes (Aidain, Espoo, Finland), with stable faecal microbiota results over multiple days when stored at RT (18–20). A change in bacterial profile was, however, demonstrated over time when the OC-sensor FIT was stored at 4°C and 30°C (21,48), and previous work has indicated that storage of stool samples at RT for four days resulted in changes of *Bifidobacterium* species, and a decrease in the relative abundance of *Faecalibacterium*, *Ruminococcus*, *Lachnospiraceae* and *Anaerostipes* (47). We did not observe such changes in the EXTEL HEMO-AUTO MC qFIT samples stored at RT for 4 days or stored at 4°C (Supplementary Figure 3), suggesting that the preservative in the EXTEL HEMO-AUTO MC qFIT buffer/faecal suspension could stabilise the overall microbiota profiles. Other previous work on the OC-sensor did support sample stability of whole stool samples at 4°C, in agreement with our findings (47,49). A limitation of our study is that we only extracted DNA at t=14 (after 4 days of storage at RT and 10 days of storage at 4^0^C). As such, we can only confirm sample stability and comparability of EXTEL HEMO-AUTO MC qFITs for 10 days post sample delivery to the laboratory.

It has previously been reported that incorrect sample collection using qFITs can alter the amount of haemoglobin measured (50), and improper sampling might explain potentially insufficient DNA recovery for 25% of the symptomatic qFITs. Gudra et al. explored lyophilisation of OC-sensor FIT samples with a low DNA amount, and found that this increased the DNA yield by up to a factor of 30, thus allowing for effective 16S rRNA gene sequencing (44). A similar approach may be worthy of further investigation for the EXTEL HEMO-AUTO MC method. Considerations regarding the gut microbiota also need to be made when sequencing symptomatic patients, as blood in the stool has been shown to influence the composition of the gut microbiota in OC-AutoFIT samples, potentially due to the varying absorption and utilisation of iron from haemoglobin by different bacterial species (51,52).

Importantly, our results indicate that the EXTEL HEMO-AUTO MC qFIT can be used for both haemoglobin screening and microbiota analysis. Given the challenges of collecting, storing, processing and comprehensively assessing the gut microbiota using larger volume/whole stool samples in both clinical and research settings, the utility of such an acceptable, easy to use, reliable and reproducible method of sampling which already has widespread clinical use cannot be understated. Additionally, as not all qFIT buffer/faecal suspension is required for 16S rRNA gene sequencing, the remainder of the buffer could be used for other analyses.

As 55% of samples from sample set three gave a DNA yield below 1 ng/µl, shotgun metagenomics-based approaches would not be able to effectively sequence the gut microbiota of most of these qFIT samples that were collected by the general population (53,54). As such, 16S rRNA gene sequencing likely remains the most practical solution to sequence these qFIT samples, albeit future shotgun metagenomic in a subset may not be precluded but will be the subject of future studies.

Several studies of OC-sensor and patient outcome data have investigated a relationship between the gut microbiota and colonoscopy results (17,43), and this would be worthy of studying with the EXTEL HEMO-AUTO MC system. Significant but subtle differences were found when comparing the microbial composition between different colonoscopy outcomes in individuals with haemoglobin positive OC-sensor FIT samples. Some 41 differentially abundant species were found when comparing colorectal cancer to non-colorectal cancer proportional abundances from the OC-sensor faecal samples of different patients (17).

Another study found an increased abundance of bacterial content in patients with high-grade dysplasia and CRC (18), and models used to distinguish patients with carcinomas from healthy patients using the gut microbiota were equally predictive using stool-derived DNA or OC-sensor FIT-derived DNA (43). The EXTEL HEMO-AUTO MC could also play an important role in established and advanced CRC given the developing evidence base relating the gut microbiome to key factors such as treatment resistance and disease recurrence (55), with samples at diagnosis and during treatment informing treatment allocation (56). Similarly, the EXTEL HEMO-AUTO MC could be employed to study microbiome-associated diseases including inflammatory bowel disease (57), cardiovascular and metabolic diseases and Alzheimer’s disease (37–40), providing timely and clinically relevant results which are easier to obtain and process than with freshly collected stool samples. As individuals are screened in the bowel screening programme routinely every two years from 50-74 years, using qFITs would also allow for longitudinal screening of the gut microbiota (10,58). More generally, longitudinal studies of healthy cohorts would also facilitate study of the effects of ageing on the gut microbiota (12,59).

Both broad range 16S rRNA gene sequencing and targeted Reverse Transcription quantitative Polymerase Chain Reaction (RT-qPCR) are currently used in clinical laboratories primarily for the accurate identification of infective pathogens (60). In the clinical setting these techniques are applied to a variety of tissue and fluid types which could extend to include leftover extraction from qFIT samples (61). Most screening and biochemistry labs will keep qFIT samples for around 14 days after analysis of f-Hb with the results of this study suggesting that this could represent a useful window for microbiota analysis. Such a resource provides a highly valuable research tool to gain greater understanding into the role, stability over time and clinical relevance of enterotypes, dominant microbiota, and functional and dysfunctional microbiomes, using large and diverse cohorts. We could then envisage these 16S rRNA gene sequencing studies identifying a useful panel of microorganisms associated with CRC, which could then be detected with high sensitivity by RT-qPCR. The cost of testing a smaller panel of bacteria by RT-qPCR in routine clinical practice could be offset by saving the high cost of colonoscopies in those without CRC, and this approach would reduce the need for unpleasant invasive investigations for patients without CRC whilst improving access to timely colonoscopy for those with CRC (62).

In conclusion, our work provides evidence for the utility of the EXTEL HEMO-AUTO MC qFIT sampling system, currently used in Scotland, Ireland, Wales, Japan and Taiwan for symptomatic testing and bowel cancer screening and in some NHS trusts in England for symptomatic testing, in determining gut microbiota profiles of individuals. This has potential to use otherwise discarded qFIT cassettes as an enormous potential resource, which could be exploited to generate highly valuable microbiome data. In the longer term, this data could be exploited to develop the prospect of future intervention studies to improve outcomes in a large range of chronic diseases linked to microbiota perturbations.

## Data Availability

The datasets generated during and/or analysed during the current study are available from the corresponding author on reasonable request.
The raw sequencing datasets used for this study have been deposited in the NCBI Short Read Archive, under accession number PRJNA1268008.

https://www.ncbi.nlm.nih.gov/bioproject/PRJNA1268008

## Conflicts of Interest

AMJ holds a voluntary committee role with the Nutrition Society, Association for the study of Obesity and British Nutrition Foundation. AWW has received research funding from ZOE Ltd, Red Rocket Labs Ltd and undertakes consultancy for EnteroBiotix Ltd. AEK has received Orafti®Synergy1 from Beneo as a generous gift for a clinical trial. The other authors declare no conflict of interest.

## Funding information

This work was funded by the NHS Grampian Endowment Fund. CRUK Scotland Centre funding enabled collaboration and research development (CTRQQR-2021\100006) and this project was supported by the RSE Research Workship grant 2399. MvdH is funded by a PhD studentship funded by the University of Aberdeen Development Trust and Friends of ANCHOR. AWW, AMJ, GH and the Rowett Institute receive core funding from the Scottish Government’s Rural and Environment Science and Analytical Services Division (RESAS). FVND’s Chair is funded by Bowel Cancer UK-Royal College of Surgeons of Edinburgh.

## Ethical approval and consent to participate

The study protocol was approved by the Rowett Ethics committee and the NHS Grampian Biorepository (TR000330), and the study was registered on clinicaltrials.gov (NCT06100549). All 16 healthy volunteer participants provided informed consent before starting the study. The two samples from patients with prostate cancer and 100 symptomatic samples were anonymously obtained under NHS Grampian Biorepository consent.

## Consent for publication

Not applicable

## Author contributions (CRediT statement)

Conceptualisation: CH, CG, AEK Patient recruitment: MAvdH and JTZ Resources: JW, ECD

Methodology and investigation: MAvdH, JTZ, ECD Data analysis and curation: MAvdH, AM, AWW, GH Writing - Original draft preparation: MAvdH

Writing - review and editing, visualisation: AEK, AMJ, CH, CG, SMcS, FD, JTZ, MAvdH, ECD, GH, AM, AWW, JW

Supervision: AEK, AMJ

Funding acquisition: AEK, CH, CG, STS, FVND

All authors reviewed the final manuscript and agreed to publication.

## Acknowledgements

We thank all volunteers that provided us with faecal samples. We thank the NHS Grampian Biorepository at Aberdeen Royal Infirmary for providing the faecal samples from cancer patients and surplus symptomatic qFIT samples, and Fiona Brandie, Katie Cameron and Julie Hawken, Biochemistry Department, Aberdeen Royal Infirmary, for their expert handling of the symptomatic qFIT samples. We thank Zeynab Heidari for her expert technical processing of the 16S rRNA gene sequencing samples. Anne Kiltie is the Friends of ANCHOR Clinical Chair in Oncology, University of Aberdeen. For the purpose of open access, the author has applied a Creative Commons Attribution (CC BY) licence to any Author Accepted Manuscript version arising from this submission.

The original version of this article has been posted as a preprint on MedRxiv: van den Haak MA, Zbikowski JT, Moomin A, Wilson J, Halsey C, Gourley C, *et al*. Accurate profiling of the gut microbiota using surplus clinical Faecal Immunochemical Test (FIT) samples. medRxiv. Preprint posted online 7 Jul 2025, https://medrxiv.org/cgi/content/short/2025.07.04.25330898v1

## Notes

### Author Declarations

The ethical committee of the Rowett Institute gave ethical approval for this work. The ethical committee of NHS Grampian Biorepository gave ethical approval for this work.

### Summary of Updates

Farhat Din changed to Farhat VN Din to include middle initials. In author contributions CG and C added to funding acquisition.

## References

1. Gerrard AD, Garau R, Xu W, Maeda Y, Dunlop MG, Theodoratou E, et al. Repeat Faecal Immunochemical Testing for Colorectal Cancer Detection in Symptomatic and Screening Patients: A Systematic Review and Meta-Analysis. Cancers (Basel). 2024 Sep 19;16(18):3199. doi:10.3390/cancers16183199 PubMed PMID: 39335170; PubMed Central PMCID: PMC11429846.

2. Cancer Research UK [Internet]. 2015 [cited 2025 Apr 24]. Bowel cancer statistics. Available from: https://www.cancerresearchuk.org/health-professional/cancer-statistics/statistics-by-cancer-type/bowel-cancer

3. Overview | Quantitative faecal immunochemical testing to guide colorectal cancer pathway referral in primary care | Guidance | NICE [Internet]. NICE; 2023 [cited 2025 Apr 24]. Available from: https://www.nice.org.uk/guidance/dg56

4. Nesci A, Carnuccio C, Ruggieri V, D’Alessandro A, Di Giorgio A, Santoro L, et al. Gut Microbiota and Cardiovascular Disease: Evidence on the Metabolic and Inflammatory Background of a Complex Relationship. Int J Mol Sci. 2023 May 22;24(10):9087. doi:10.3390/ijms24109087 PubMed PMID: 37240434; PubMed Central PMCID: PMC10219307.

5. Qiu P, Ishimoto T, Fu L, Zhang J, Zhang Z, Liu Y. The Gut Microbiota in Inflammatory Bowel Disease. Front Cell Infect Microbiol. 2022 Feb 22;12:733992. doi:10.3389/fcimb.2022.733992 PubMed PMID: 35273921; PubMed Central PMCID: PMC8902753.

6. Kim J, Lee HK. Potential Role of the Gut Microbiome In Colorectal Cancer Progression. Front Immunol. 2022 Jan 7;12:807648. doi:10.3389/fimmu.2021.807648 PubMed PMID: 35069592; PubMed Central PMCID: PMC8777015.

7. Johnson JS, Spakowicz DJ, Hong BY, Petersen LM, Demkowicz P, Chen L, et al. Evaluation of 16S rRNA gene sequencing for species and strain-level microbiome analysis. Nat Commun. 2019 Nov 6;10(1):5029. doi:10.1038/s41467-019-13036-1

8. Abellan-Schneyder I, Matchado MS, Reitmeier S, Sommer A, Sewald Z, Baumbach J, et al. Primer, Pipelines, Parameters: Issues in 16S rRNA Gene Sequencing. mSphere. 6(1):e01202–20. doi:10.1128/mSphere.01202-20 PubMed PMID: 33627512; PubMed Central PMCID: PMC8544895.

9. NHS Research Scotland | NHS Research Scotland [Internet]. [cited 2025 Mar 28]. Available from: https://www.nhsresearchscotland.org.uk/research-in-scotland/data/safe-havens

10. Björk JR, Bolte LA, Maltez Thomas A, Lee KA, Rossi N, Wind TT, et al. Longitudinal gut microbiome changes in immune checkpoint blockade-treated advanced melanoma. Nat Med. 2024 Mar;30(3):785–96. doi:10.1038/s41591-024-02803-3

11. Rezasoltani S, Azizmohammad Looha M, Asadzadeh Aghdaei H, Jasemi S, Sechi LA, Gazouli M, et al. 16S rRNA sequencing analysis of the oral and fecal microbiota in colorectal cancer positives versus colorectal cancer negatives in Iranian population. Gut Pathogens. 2024 Feb 20;16(1):9. doi:10.1186/s13099-024-00604-0

12. Ragonnaud E, Biragyn A. Gut microbiota as the key controllers of “healthy” aging of elderly people. Immunity & Ageing. 2021 Jan 5;18(1):2. doi:10.1186/s12979-020-00213-w

13. Inamura K, Hamada T, Bullman S, Ugai T, Yachida S, Ogino S. Cancer as microenvironmental, systemic, and environmental diseases: opportunity for transdisciplinary microbiomics science. Gut. 2022 Jul 12;gutjnl-2022-327209. doi:10.1136/gutjnl-2022-327209 PubMed PMID: 35820782; PubMed Central PMCID: PMC9834441.

14. Georgiou Delisle T, D’Souza N, Davies B, Benton S, Chen M, Ward H, et al. Faecal immunochemical test for suspected colorectal cancer symptoms: patient survey of usability and acceptability. BJGP Open. 2022 Mar;6(1):BJGPO.2021.0102. doi:10.3399/BJGPO.2021.0102 PubMed PMID: 34645655; PubMed Central PMCID: PMC8958743.

15. Baxter NT, Koumpouras CC, Rogers MAM, Ruffin MT, Schloss PD. DNA from fecal immunochemical test can replace stool for detection of colonic lesions using a microbiota-based model. Microbiome. 2016 Nov 14;4:59. doi:10.1186/s40168-016-0205-y PubMed PMID: 27842559; PubMed Central PMCID: PMC5109736.

16. Rounge TB, Meisal R, Nordby JI, Ambur OH, de Lange T, Hoff G. Evaluating gut microbiota profiles from archived fecal samples. BMC Gastroenterol. 2018 Nov 8;18:171. doi:10.1186/s12876-018-0896-6 PubMed PMID: 30409123; PubMed Central PMCID: PMC6225565.

17. Khannous-Lleiffe O, Willis JR, Saus E, Moreno V, Castellví-Bel S, Gabaldón T. Microbiome Profiling from Fecal Immunochemical Test Reveals Microbial Signatures with Potential for Colorectal Cancer Screening. Cancers (Basel). 2022 Dec 25;15(1):120. doi:10.3390/cancers15010120 PubMed PMID: 36612118; PubMed Central PMCID: PMC9817783.

18. Grobbee EJ, Lam SY, Fuhler GM, Blakaj B, Konstantinov SR, Bruno MJ, et al. First steps towards combining faecal immunochemical testing with the gut microbiome in colorectal cancer screening. United European Gastroenterol J. 2020 Apr;8(3):293–302. doi:10.1177/2050640619890732 PubMed PMID: 32213018; PubMed Central PMCID: PMC7184657.

19. Vogtmann E, Chen J, Amir A, Shi J, Abnet CC, Nelson H, et al. Comparison of Collection Methods for Fecal Samples in Microbiome Studies. Am J Epidemiol. 2017 Jan 15;185(2):115–23. doi:10.1093/aje/kww177 PubMed PMID: 27986704; PubMed Central PMCID: PMC5253972.

20. Krigul KL, Aasmets O, Lüll K, Org T, Org E. Using fecal immunochemical tubes for the analysis of the gut microbiome has the potential to improve colorectal cancer screening. Sci Rep. 2021 Oct 1;11(1):19603. doi:10.1038/s41598-021-99046-w

21. Zouiouich S, Mariadassou M, Rué O, Vogtmann E, Huybrechts I, Severi G, et al. Comparison of fecal sample collection methods for microbial analysis embedded within colorectal cancer screening programs. Cancer Epidemiol Biomarkers Prev. 2022 Feb;31(2):305–14. doi:10.1158/1055-9965.EPI-21-0188 PubMed PMID: 34782392; PubMed Central PMCID: PMC10416615.

22. QuikRead go iFOBT: Quantitative iFOBT (FIT) test | Aidian [Internet]. [cited 2025 Oct 16]. Available from: https://www.aidian.eu/point-of-care/quikread-go/quikread-go-ifobt#documents-and-materials

23. Fraser CG, Allison JE, Halloran SP, Young GP, Expert Working Group on Fecal Immunochemical Tests for Hemoglobin, Colorectal Cancer Screening Committee, World Endoscopy Organization. A proposal to standardize reporting units for fecal immunochemical tests for hemoglobin. J Natl Cancer Inst. 2012 Jun 6;104(11):810–4. doi:10.1093/jnci/djs190 PubMed PMID: 22472305.

24. Faecal immunochemical test sample Collection Picker. To collect, preserve, and test samples for analysis on the HM-JACKarc analyser. Pack of 200. [Internet]. [cited 2024 Jul 31]. Available from: https://www.alphalabs.co.uk/063631

25. FIT Screening - UK - FAQs [Internet]. [cited 2024 Jul 31]. Available from: http://www.fit-screening.co.uk/resources/faqs

26. Cancer Research UK [Internet]. 2023 [cited 2025 Apr 28]. A study looking at how bacteria in the gut affects treatment for pelvic cancer (PELICAN-23). Available from: https://www.cancerresearchuk.org/about-cancer/find-a-clinical-trial/a-study-looking-at-how-bacteria-in-the-gut-affects-treatment-for-pelvic-cancer-pelican-23

27. NHS research scotland [Internet]. [cited 2025 Oct 9]. Available from: https://www.nhsresearchscotland.org.uk/research-in-scotland/facilities/biorepositories-and-tissue-services/overview

28. Then CK, Paillas S, Moomin A, Misheva MD, Moir RA, Hay SM, et al. Dietary fibre supplementation enhances radiotherapy tumour control and alleviates intestinal radiation toxicity. Microbiome. 2024 May 14;12:89. doi:10.1186/s40168-024-01804-1 PubMed PMID: 38745230; PubMed Central PMCID: PMC11092108.

29. Mukhopadhya I, Martin JC, Shaw S, McKinley AJ, Gratz SW, Scott KP. Comparison of microbial signatures between paired faecal and rectal biopsy samples from healthy volunteers using next-generation sequencing and culturomics. Microbiome. 2022 Oct 14;10(1):171. doi:10.1186/s40168-022-01354-4

30. Schloss PD, Westcott SL, Ryabin T, Hall JR, Hartmann M, Hollister EB, et al. Introducing mothur: Open-Source, Platform-Independent, Community-Supported Software for Describing and Comparing Microbial Communities. Appl Environ Microbiol. 2009 Dec;75(23):7537–41. doi:10.1128/AEM.01541-09 PubMed PMID: 19801464; PubMed Central PMCID: PMC2786419.

31. Kozich JJ, Westcott SL, Baxter NT, Highlander SK, Schloss PD. Development of a Dual-Index Sequencing Strategy and Curation Pipeline for Analyzing Amplicon Sequence Data on the MiSeq Illumina Sequencing Platform. Applied and Environmental Microbiology. 2013 Sep;79(17):5112–20. doi:10.1128/AEM.01043-13

32. Huse SM, Welch DM, Morrison HG, Sogin ML. Ironing out the wrinkles in the rare biosphere through improved OTU clustering. Environ Microbiol. 2010 Jul;12(7):1889–98. doi:10.1111/j.1462-2920.2010.02193.x PubMed PMID: 20236171; PubMed Central PMCID: PMC2909393.

33. Wang Q, Garrity GM, Tiedje JM, Cole JR. Naive Bayesian classifier for rapid assignment of rRNA sequences into the new bacterial taxonomy. Appl Environ Microbiol. 2007 Aug;73(16):5261–7. doi:10.1128/AEM.00062-07 PubMed PMID: 17586664; PubMed Central PMCID: PMC1950982.

34. White JR, Nagarajan N, Pop M. Statistical methods for detecting differentially abundant features in clinical metagenomic samples. PLoS Comput Biol. 2009 Apr;5(4):e1000352. doi:10.1371/journal.pcbi.1000352 PubMed PMID: 19360128; PubMed Central PMCID: PMC2661018.

35. Benjamini Y, Hochberg Y. Controlling the False Discovery Rate: A Practical and Powerful Approach to Multiple Testing. Journal of the Royal Statistical Society: Series B (Methodological). 1995;57(1):289–300. doi:10.1111/j.2517-6161.1995.tb02031.x

36. Nearing JT, Douglas GM, Hayes MG, MacDonald J, Desai DK, Allward N, et al. Microbiome differential abundance methods produce different results across 38 datasets. Nat Commun. 2022 Jan 17;13(1):342. doi:10.1038/s41467-022-28034-z

37. Segata N, Izard J, Waldron L, Gevers D, Miropolsky L, Garrett WS, et al. Metagenomic biomarker discovery and explanation. Genome Biology. 2011 Jun 24;12(6):R60. doi:10.1186/gb-2011-12-6-r60

38. Chong J, Liu P, Zhou G, Xia J. Using MicrobiomeAnalyst for comprehensive statistical, functional, and meta-analysis of microbiome data. Nat Protoc. 2020 Mar;15(3):799–821. doi:10.1038/s41596-019-0264-1

39. Finotello F, Mastrorilli E, Di Camillo B. Measuring the diversity of the human microbiota with targeted next-generation sequencing. Briefings in Bioinformatics. 2018 Jul 20;19(4):679–92. doi:10.1093/bib/bbw119

40. Letunic I, Bork P. Interactive Tree of Life (iTOL) v6: recent updates to the phylogenetic tree display and annotation tool. Nucleic Acids Research. 2024 Jul 5;52(W1):W78–82. doi:10.1093/nar/gkae268

41. Schloss PD, Handelsman J. Introducing TreeClimber, a test to compare microbial community structures. Appl Environ Microbiol. 2006 Apr;72(4):2379–84. doi:10.1128/AEM.72.4.2379-2384.2006 PubMed PMID: 16597933; PubMed Central PMCID: PMC1449046.

42. Mallick H, Rahnavard A, McIver LJ, Ma S, Zhang Y, Nguyen LH, et al. Multivariable association discovery in population-scale meta-omics studies. PLOS Computational Biology. 2021 Nov 16;17(11):e1009442. doi:10.1371/journal.pcbi.1009442

43. Baxter NT, Koumpouras CC, Rogers MAM, Ruffin MT, Schloss PD. DNA from fecal immunochemical test can replace stool for detection of colonic lesions using a microbiota-based model. Microbiome. 2016 Nov 14;4(1):59. doi:10.1186/s40168-016-0205-y PubMed PMID: 27842559; PubMed Central PMCID: PMC5109736.

44. Gudra D, Shoaie S, Fridmanis D, Klovins J, Wefer H, Silamikelis I, et al. A widely used sampling device in colorectal cancer screening programmes allows for large-scale microbiome studies. Gut. 2019 Sep;68(9):1723–5. doi:10.1136/gutjnl-2018-316225 PubMed PMID: 30242040; PubMed Central PMCID: PMC6709769.

45. Brezina S, Borkovec M, Baierl A, Bastian F, Futschik A, Gasche N, et al. Using fecal immmunochemical cartridges for gut microbiome analysis within a colorectal cancer screening program. Gut Microbes. 15(1):2176119. doi:10.1080/19490976.2023.2176119 PubMed PMID: 36794815; PubMed Central PMCID: PMC9980522.

46. Byrd DA, Chen J, Vogtmann E, Hullings A, Song SJ, Amir A, et al. Reproducibility, stability, and accuracy of microbial profiles by fecal sample collection method in three distinct populations. PLoS One. 2019 Nov 18;14(11):e0224757. doi:10.1371/journal.pone.0224757 PubMed PMID: 31738775; PubMed Central PMCID: PMC6860998.

47. Choo JM, Leong LE, Rogers GB. Sample storage conditions significantly influence faecal microbiome profiles. Sci Rep. 2015 Nov 17;5:16350. doi:10.1038/srep16350 PubMed PMID: 26572876; PubMed Central PMCID: PMC4648095.

48. Birkeland E, Ferrero G, Pardini B, Umu SU, Tarallo S, Bulfamante S, et al. Profiling small RNAs in fecal immunochemical tests: is it possible? Mol Cancer. 2023 Oct 3;22(1):161. doi:10.1186/s12943-023-01869-w

49. Holzhausen EA, Nikodemova M, Deblois CL, Barnet JH, Peppard PE, Suen G, et al. Assessing the impact of storage time on the stability of stool microbiota richness, diversity, and composition. Gut Pathog. 2021 Dec 20;13:75. doi:10.1186/s13099-021-00470-0 PubMed PMID: 34930464; PubMed Central PMCID: PMC8686582.

50. Gies A, Gruner LF, Schrotz-King P, Brenner H. Effect of Imperfect Compliance With Instructions for Fecal Sample Collection on Diagnostic Performance of 9 Fecal Immunochemical Tests. Clinical Gastroenterology and Hepatology. 2019 Aug 1;17(9):1829–1839.e4. doi:10.1016/j.cgh.2019.03.001

51. Chénard T, Malick M, Dubé J, Massé E. The influence of blood on the human gut microbiome. BMC Microbiol. 2020 Mar 3;20:44. doi:10.1186/s12866-020-01724-8 PubMed PMID: 32126968; PubMed Central PMCID: PMC7055051.

52. Seyoum Y, Baye, Kaleab, and Humblot C. Iron homeostasis in host and gut bacteria – a complex interrelationship. Gut Microbes. 2021 Jan 1;13(1):1874855. doi:10.1080/19490976.2021.1874855 PubMed PMID: 33541211.

53. Shotgun Metagenomic Sequencing. Novogene [Internet]. [cited 2025 Dec 18]. Available from: https://www.novogene.com/eu-en/services/research-services/metagenome-sequencing/shotgun-metagenomic-sequencing/

54. Zymo Research International [Internet]. [cited 2025 Dec 18]. Shotgun Metagenomic Sequencing Service | ZYMO RESEARCH. Available from: https://zymoresearch.eu/pages/shotgun-metagenomic-sequencing-service

55. Teng H, Wang Y, Sui X, Fan J, Li S, Lei X, et al. Gut microbiota-mediated nucleotide synthesis attenuates the response to neoadjuvant chemoradiotherapy in rectal cancer. Cancer Cell. 2023 Jan 9;41(1):124–138.e6. doi:10.1016/j.ccell.2022.11.013 PubMed PMID: 36563680.

56. Cheng WY, Wu CY, Yu J. The role of gut microbiota in cancer treatment: friend or foe? Gut. 2020 Oct;69(10):1867–76. doi:10.1136/gutjnl-2020-321153

57. Bethlehem L, Estevinho MM, Grinspan A, Magro F, Faith JJ, Colombel JF. Microbiota therapeutics for inflammatory bowel disease: the way forward. Lancet Gastroenterol Hepatol. 2024 May;9(5):476–86. doi:10.1016/S2468-1253(23)00441-7 PubMed PMID: 38604201.

58. Olsson LM, Boulund F, Nilsson S, Khan MT, Gummesson A, Fagerberg L, et al. Dynamics of the normal gut microbiota: A longitudinal one-year population study in Sweden. Cell Host & Microbe. 2022 May 11;30(5):726–739.e3. doi:10.1016/j.chom.2022.03.002

59. Ghosh TS, Shanahan F, O’Toole PW. The gut microbiome as a modulator of healthy ageing. Nat Rev Gastroenterol Hepatol. 2022 Sep;19(9):565–84. doi:10.1038/s41575-022-00605-x

60. Yuen W. Then and now: use of 16S rDNA gene sequencing for bacterial identification and discovery of novel bacteria in clinical microbiology laboratories. Clinical Microbiology and Infection. 2008 Oct 1;14(10):908–34. doi:10.1111/j.1469-0691.2008.02070.x

61. Harris KA, Brown JR. Diagnostic yield of broad-range 16s rRNA gene PCR varies by sample type and is improved by the addition of qPCR panels targeting the most common causative organisms. Journal of Medical Microbiology. 2022;71(12):001633. doi:10.1099/jmm.0.001633

62. Re-evaluating post-polypectomy surveillance: The role of non-invasive modalities in colorectal cancer prevention. Best Practice & Research Clinical Gastroenterology. 2026 Feb 1;80:102092.doi:10.1016/j.bpg.2026.102092

