## Supplemental Figure 1 for "Reproducible profiling of the gut microbiota using surplus clinical Faecal Immunochemical Test (FIT) samples"

### Supplementary figures

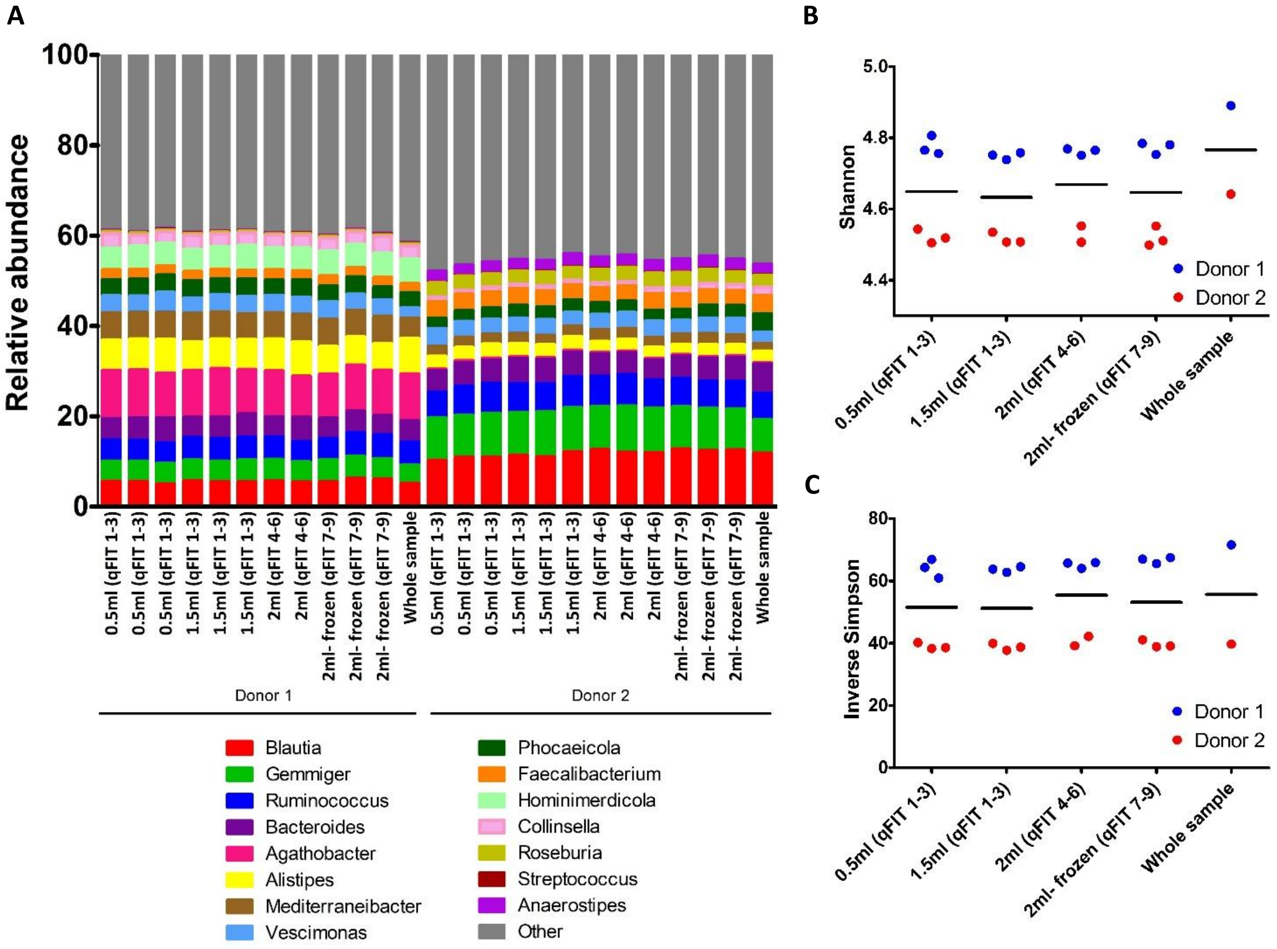


**Figure S1. qFIT sampling volume did not influence bacterial proportional abundance and alpha diversity measures.** A) Relative abundance data for donor 1 and donor 2 at the genus level. Metastats on the 100 most proportionally abundant OTU's and phylotypes with Benjamini-Hochberg correction and Benjamini-Hochberg adjusted LEfSe and DESeq2 were conducted to evaluate statistically significant differences; B) Shannon diversity index, separated by donor 1 and 2, with the bar representing the overall mean between the two donors. ANOVA was used to assess the significance of differences among the five experimental conditions; C) Inverse Simpson index scores, with samples separated by donor 1 and 2. ANOVA was applied to evaluate statistical differences among the experimental conditions.

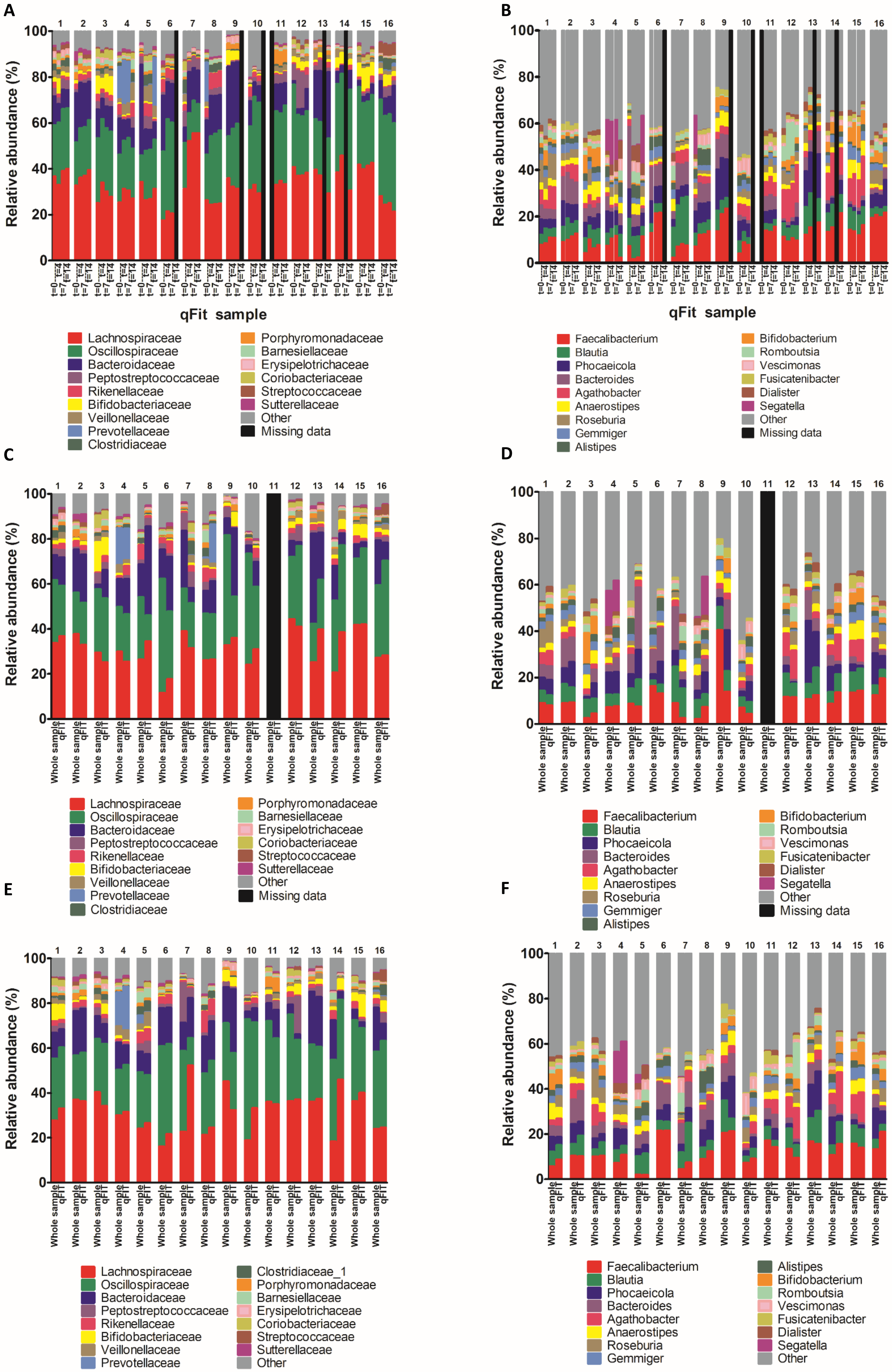
A
B
C
D
E
F
Relative abundance (%)
qFit sample
t=0
t=7
t=14
Whole sample
qFit
Lachnospiraceae
Oscillospiraceae
Bacteroidaceae
Peptostreptococcaceae
Rikenellaceae
Bifidobacteriaceae
Veillonellaceae
Prevotellaceae
Clostridiaceae
Clostridiaceae_1
Porphyromonadaceae
Barnesiellaceae
Erysipelotrichaceae
Coriobacteriaceae
Streptococcaceae
Sutterellaceae
Other
Missing data
Faecalibacterium
Blautia
Phocaeicola
Bacteroides
Agathobacter
Anaerostipes
Roseburia
Gemmiger
Alistipes
Bifidobacterium
Romboutsia
Vescimonas
Fusicatenibacter
Dialister
Segatella

**Figure S2. qFIT samples remained stable over time and showed similar microbiota profile when compared to whole stool-derived samples at the family and genus levels.** Metastats on the 100 most proportionally abundant OTU's and phylotypes with Benjamini-Hochberg correction and Benjamini-Hochberg adjusted LEfSe and DESeq2 were conducted to evaluate statistically significant differences. A) relative bacterial abundances for qFIT over time at the family level and B) genus level. C) Relative bacterial abundance for qFIT and whole stool sample at t=0 at the family level, and D) genus level. E) Relative bacterial abundance of qFIT and whole stool sample at t=4 at the family and F) genus level.
